# Who Gets a Flu Shot?

**DOI:** 10.64898/2025.12.03.25341550

**Authors:** James McIntosh

## Abstract

**Objective:** To determine the characteristics which lead individuals to get a flu shot.

**Study Design:** Two countries are examined using sample survey data to compare the relative contribution of education and self-perceived health in the flu shot decision.

**Methods:** Logit probability models are fitted to explain the binary decision to have a flu shot using the Canadian Covid-19 Antibody Survey for 2020 and the American National Health Interview Survey for 2022. Regressors included age gender, self reported health status and educational attainment.

**Results:** Educational attainment was shown to be the main driver of this decision in the US whereas self reported health explained most of the variation in the Canadian data. The difference in the results was attributed to differences in the health care systems.

**Conclusions:** Both countries have unacceptably low vaccination rates for influenza. For the US public policies should do a better job in addressing this problem by concentrating efforts on getting more low income or poorly educated respondents to get vaccinated. For Canada, the un-vaxinated are less visible but public education programs could be improved.

## 1 Introduction

### 1.1 Background

Flu or influenza as it is known in medical circles is a serious illness. In both Canada and the United States, the two countries considered in this paper, it ranks as the 8^th^ or 9^th^ largest cause of death (Health Canada (2023) and CDC (2023))^1^. Between 5 and 10 percent of adults and 20 to 30 percent of children are affected each year. Both countries combine flu data with pneumonia as a category since about 27% of influenza cases also suffer from pneumonia (Shoar et al (2020)). In spite of the health risks associated with flu many Canadians and Americans do not get vaccinated. Vaccination rates in the two countries are similar: 43% for Canada and 44% for the United States. The seriousness of this shortfall becomes apparent when vaccine efficacy rates are considered. American flu vaccines are only moderately effective at 38% (62% against H1N1 and 22% against H3N2) and require constant updating^2^ so this means that herd immunity can be reached only when about 80% of the population is vaccinated. This is the target set by the two monitoring health agencies but so far it has not been reached.

The low rates of participation in vaccination programs is a serious policy failure in both countries. Vaccination policy needs to be better focused and more directed towards groups who do not get vaccinated. Health researchers should examine who gets vaccinated and why large segments of both populations do not consider flu vaccines to be important. Both Health Canada and the CDC collect sample data on whether respondents have had a flu vaccine together with social and demographic characteristics but only Health Canada collects information on why respondents have or have not had a vaccine^3^.

Recent research has attempted to identify social or demographic groups with low participation rates. Watson and Oancea (2020) following the work of La et al (2018) showed using data on self reported health from the 2017 US Behavioral Risk Factor Surveillance System (BRFSS) survey that individuals with better self-reported health were less likely to get vaccinated. This result was confirmed for Canada using data from the 2017 Canadian Community Health Survey (CCHS).The most common reason given (36%) was that respondents did not think it was necessary

A number of issues are considered here. The authors suggested that their research should be extended to see whether their results hold for the US over time and ii) and to see if they hold for other countries. These two issues are examined by looking at health data for 2017, 2020, and 2022 and two countries: Canada and the United States. The second deals with some important methodological and statistical issues that arise in their paper. The third addresses the question of whether the variable, self-reported health, is the most suitable variable over which to measure variation in flu vaccination rates. The most important result obtained here is that self-reported health is not the best variable to use for analyzing the variation in vaccination rates. Levels of education or household income show much more variation in vaccination rates; they are predetermined; and it is much easier to identify respondents with the different levels of these two variables than it is for self-reported health. As will be discussed below there are other reasons for preferring this option.

## 2. Data and Methods

### 2.1 Data

The surveys are used in the analysis for the US are the Behavioral Risk Factor Surveillance System (BRFSS) 2022 and the National Health Interview Survey (NHIS) 2022. For Canada they are the Canadian Community Health Survey (CCHS) 2017 and Canadian COVID-19 Antibody and Health Survey (CCAHS) 2020. The details concerning the variables included and sample sizes are in Tables 1 and 2. Both country surveys ask respondents whether they had a flu shot in the 12 months prior to interview. There is also quite a large selection of variables which describe the characteristics of the respondent. The variables included here are age, gender, the highest of education in the household, sometimes income and race. Different surveys pose survey questions differently. The CCAHS and the NHIS have more detailed information on education than the other two surveys so they are used instead of the 2017 CCHS and the 2022 BRFSS surveys in the statistical analysis but information from other surveys is also used. Means and standard deviations for the variables appear in Tables 1 and 2.

**Table 1.**
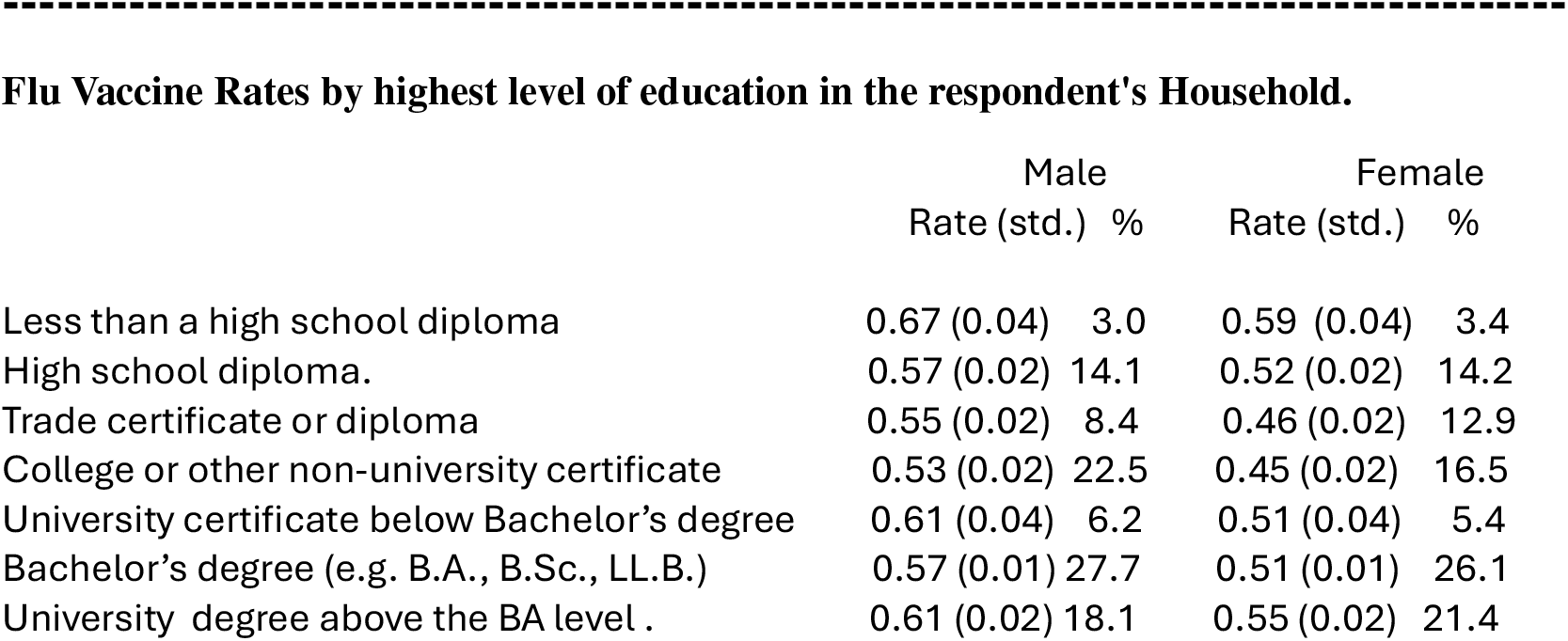
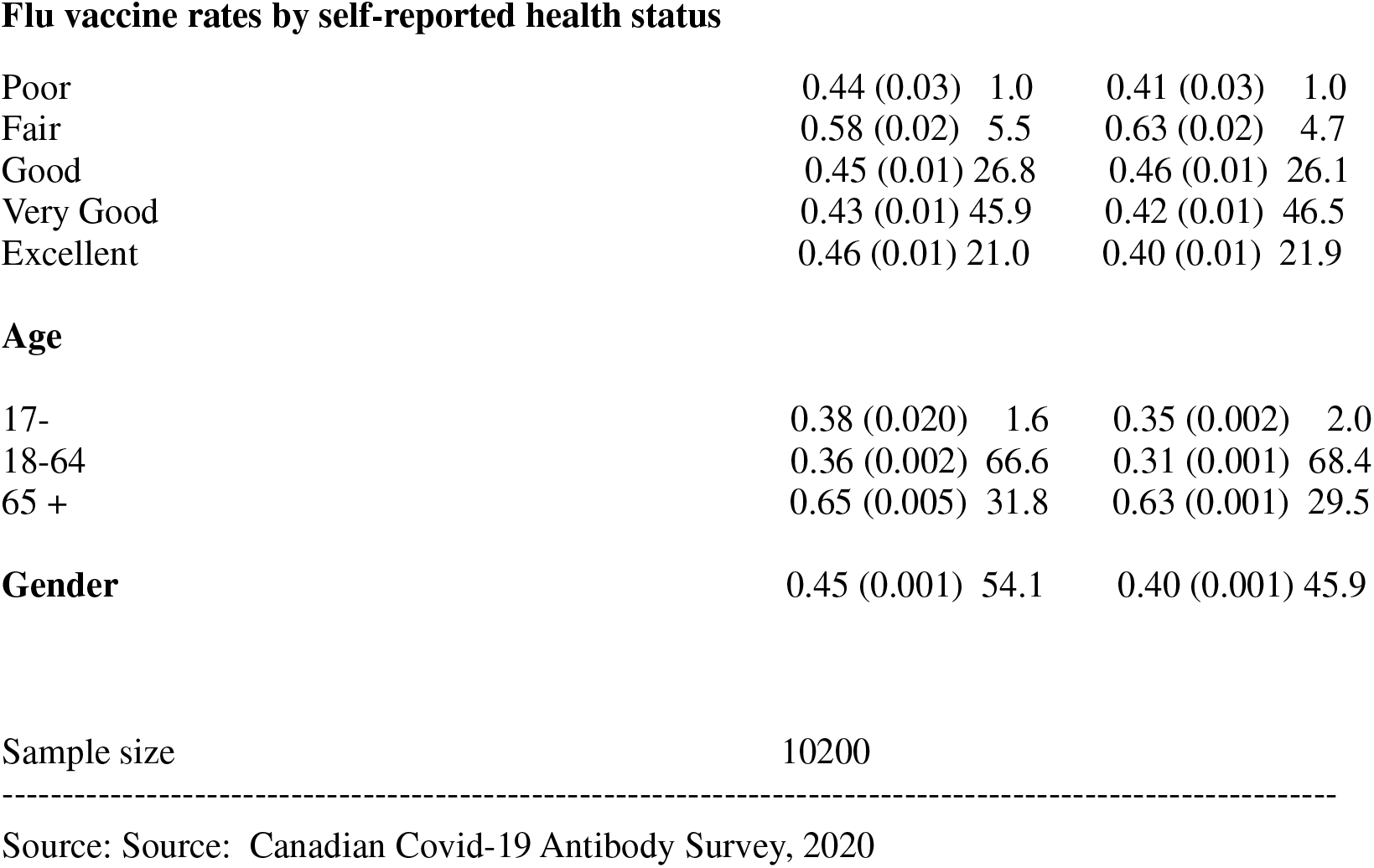
Canada. Vaccination rates by educational level, self-reported health, age, and gender.

**Flu Vaccine Rates by highest level of education in the respondent's Household.**
|  | Male |  | Female |  |
| --- | --- | --- | --- | --- |
|  | Rate (std.) | % | Rate (std.) | % |
| Less than a high school diploma | 0.67 (0.04) | 3.0 | 0.59 (0.04) | 3.4 |
| High school diploma. | 0.57 (0.02) | 14.1 | 0.52 (0.02) | 14.2 |
| Trade certificate or diploma | 0.55 (0.02) | 8.4 | 0.46 (0.02) | 12.9 |
| College or other non-university certificate | 0.53 (0.02) | 22.5 | 0.45 (0.02) | 16.5 |
| University certificate below Bachelor's degree | 0.61 (0.04) | 6.2 | 0.51 (0.04) | 5.4 |
| Bachelor's degree (e.g. B.A., B.Sc., LL.B.) | 0.57 (0.01) | 27.7 | 0.51 (0.01) | 26.1 |
| University degree above the BA level . | 0.61 (0.02) | 18.1 | 0.55 (0.02) | 21.4 |
| Flu vaccine rates by self-reported health status |  |  |  |  |
| Poor | 0.44 (0.03) | 1.0 | 0.41 (0.03) | 1.0 |
| Fair | 0.58 (0.02) | 5.5 | 0.63 (0.02) | 4.7 |
| Good | 0.45 (0.01) | 26.8 | 0.46 (0.01) | 26.1 |
| Very Good | 0.43 (0.01) | 45.9 | 0.42 (0.01) | 46.5 |
| Excellent | 0.46 (0.01) | 21.0 | 0.40 (0.01) | 21.9 |
| Age |  |  |  |  |
| 17- | 0.38 (0.020) | 1.6 | 0.35 (0.002) | 2.0 |
| 18-64 | 0.36 (0.002) | 66.6 | 0.31 (0.001) | 68.4 |
| 65 + | 0.65 (0.005) | 31.8 | 0.63 (0.001) | 29.5 |
| **Gender** | 0.45 (0.001) | 54.1 | 0.40 (0.001) | 45.9 |

Source: Source: Canadian Covid-19 Antibody Survey, 2020

**Table 2.**
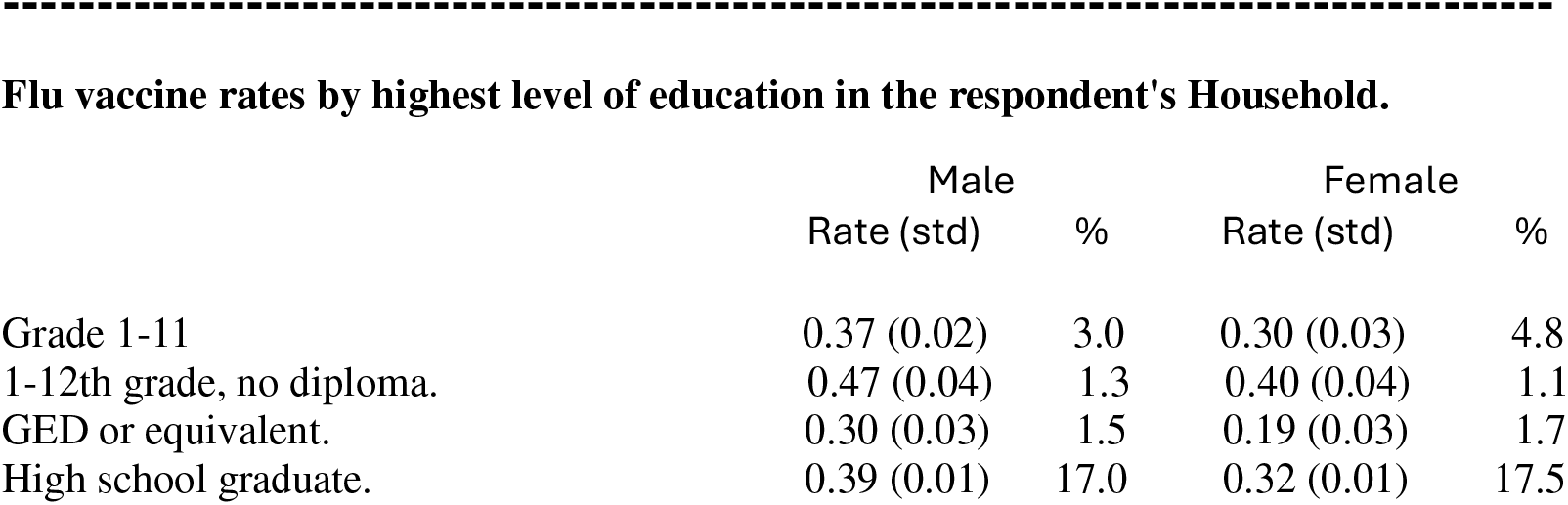
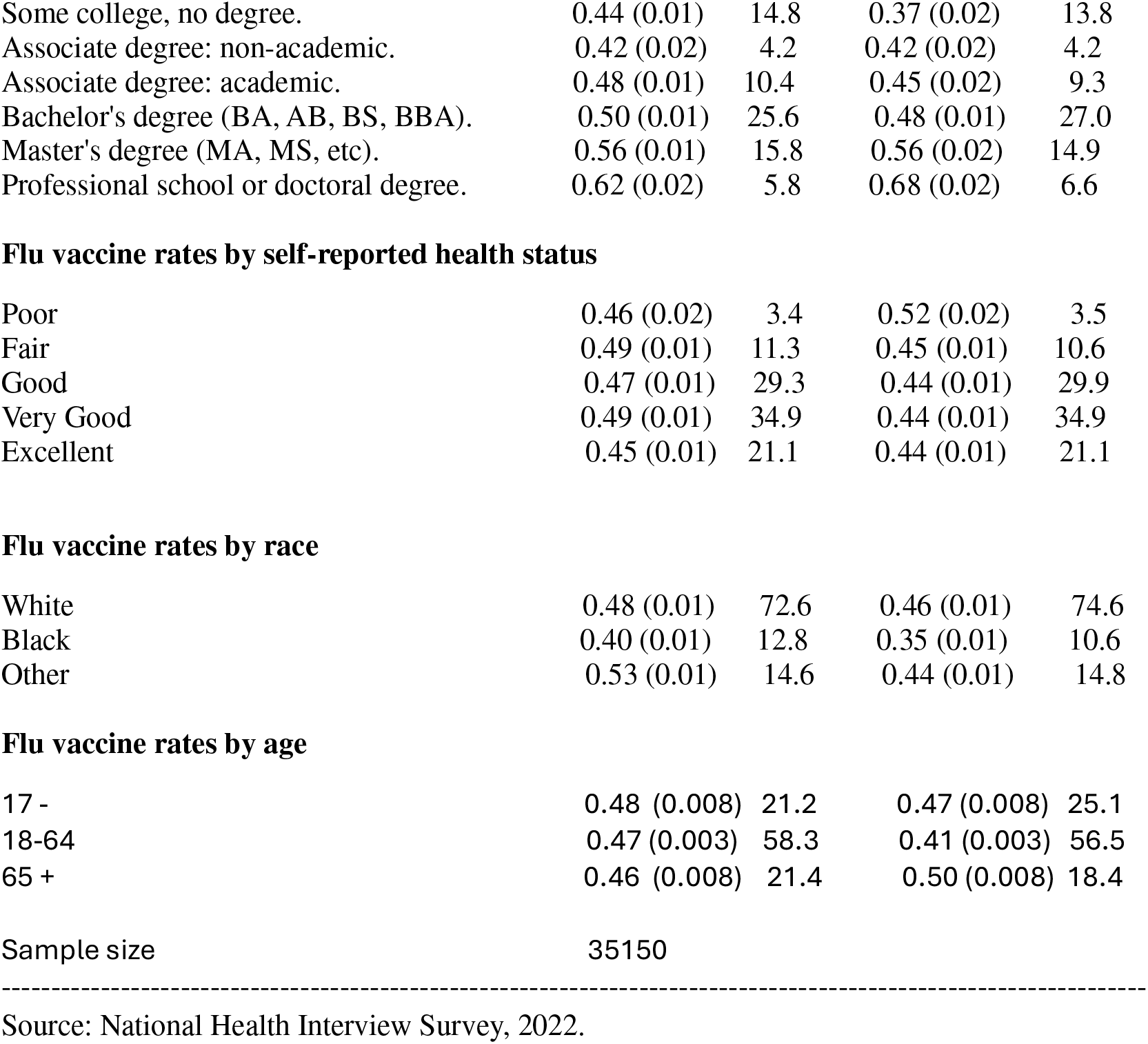
United States. Vaccination rates by educational level, self-reported health, race, age, and gender.

Flu vaccine rates by highest level of education in the respondent's Household.
|  | Male |  | Female |  |
| --- | --- | --- | --- | --- |
|  | Rate (std) | % | Rate (std) | % |
| Grade 1-11 | 0.37 (0.02) | 3.0 | 0.30 (0.03) | 4.8 |
| 1-12th grade, no diploma. | 0.47 (0.04) | 1.3 | 0.40 (0.04) | 1.1 |
| GED or equivalent. | 0.30 (0.03) | 1.5 | 0.19 (0.03) | 1.7 |
| High school graduate. | 0.39 (0.01) | 17.0 | 0.32 (0.01) | 17.5 |
| Some college, no degree. | 0.44 (0.01) | 14.8 | 0.37 (0.02) | 13.8 |
| Associate degree: non-academic. | 0.42 (0.02) | 4.2 | 0.42 (0.02) | 4.2 |
| Associate degree: academic. | 0.48 (0.01) | 10.4 | 0.45 (0.02) | 9.3 |
| Bachelor's degree (BA, AB, BS, BBA). | 0.50 (0.01) | 25.6 | 0.48 (0.01) | 27.0 |
| Master's degree (MA, MS, etc). | 0.56 (0.01) | 15.8 | 0.56 (0.02) | 14.9 |
| Professional school or doctoral degree. | 0.62 (0.02) | 5.8 | 0.68 (0.02) | 6.6 |
| Flu vaccine rates by self-reported health status |  |  |  |  |
| Poor | 0.46 (0.02) | 3.4 | 0.52 (0.02) | 3.5 |
| Fair | 0.49 (0.01) | 11.3 | 0.45 (0.01) | 10.6 |
| Good | 0.47 (0.01) | 29.3 | 0.44 (0.01) | 29.9 |
| Very Good | 0.49 (0.01) | 34.9 | 0.44 (0.01) | 34.9 |
| Excellent | 0.45 (0.01) | 21.1 | 0.44 (0.01) | 21.1 |
| Flu vaccine rates by race |  |  |  |  |
| White | 0.48 (0.01) | 72.6 | 0.46 (0.01) | 74.6 |
| Black | 0.40 (0.01) | 12.8 | 0.35 (0.01) | 10.6 |
| Other | 0.53 (0.01) | 14.6 | 0.44 (0.01) | 14.8 |
| Flu vaccine rates by age |  |  |  |  |
| 17 - | 0.48 (0.008) | 21.2 | 0.47 (0.008) | 25.1 |
| 18-64 | 0.47 (0.003) | 58.3 | 0.41 (0.003) | 56.5 |
| 65 + | 0.46 (0.008) | 21.4 | 0.50 (0.008) | 18.4 |

Sample size 35150
Source: National Health Interview Survey, 2022.

### 2.2 Statistical methodology

The variable of interest is whether respondent j received a flu vaccination in the period. Define this event as v_j_=1 for respondent j. If this binary outcome variable follows a logistic distribution then

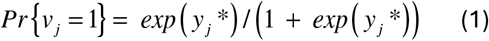

where the latent variable

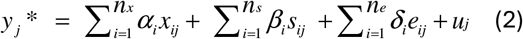

The x_ij_ variables are various regressors like age, race etc; s_ij_ are the four self-reported health dummy variables and e_ij_ are the dummy variables for the education categories listed in tables 1 and 2. The (α, β, δ) coefficients can be estimated by maximum likelihood or non-linear least squares methods.

The most interesting information about the model is contained in odds ratios. These are computed for both self-reported and educational categories in table 3 for each country. Odds are specific to each category. For self-reported health the odds for respondent j are

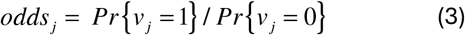

which has 5 values depending on the self-reported health category. The odds ratio for self-reported health category k for respondent j is

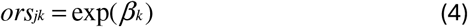

**Table 3.** Canada Odds ratios by educational level and health status.

| **Odds ratios by education level** | **Males** |  | **Females** |  |
| --- | --- | --- | --- | --- |
| Less than High School diploma | 1.0 | - | 1.0 |  |
| High school diploma. | 1.029 | (0.30) | 0.856 | (0.25) |
| Trade certificate or diploma | 0.869 | (0.26) | 0.890 | (0.26) |
| College/CEGEP/other non-university certificate | 0.890 | (0.26) | 1.080 | (0.35) |
| University certificate below the bachelor's level. | 1.099 | (0.38) | 1.220 | (0.36) |
| Bachelor's degree (e.g. B.A., B.Sc., LL.B.) | 1.391 | (0.41) | 1.368 | (0.41) |
| University degree above the BA level . | 0.61 | (0.02) | 1.813 | (0.55) |

| **Odds ratios by health status** | **Males** |  | **Females** |  |
| --- | --- | --- | --- | --- |
| Poor | 1.700 * | ( 0.15) | 1.396* | (0.10) |
| Fair | 1.365 * | ( 0.08) | 1.414* | (0.07) |
| Good | 1.084 * | ( 0.04 ) | 1.088* | (0.04) |
| Very Good | 0.955 | ( 0.03 ) | 1.111* | (0.04) |
| Excellent | 1.0 | - | 1.0 | - |

Source: Canadian Covid-19 Antibody Survey, 2020. \* means significant at least at the 5% level

These are the ratios of the category odds where category 5 (excellent health) is the residual category for self-reported health and category 1 (less than high school) is the residual category for the education variables. Because the logistic distribution is being used odds ratios do not depend on regressors and are the same for all respondents. This is not a characteristic of other distributions. Later it will be shown that other distributions explain the data equally well.

## 3. Results

The first point to note is that the result that Watson and Oancea found is confirmed for all of the surveys considered in his paper. The American distribution of self-reported health outcomes are almost identical for the 2017 and 2022 BRFSS surveys. This means that Covid 19 had no impact on American health outcomes over the five years between the two surveys. Results are similar for Canada. Both the CCHS and CCAHS show that respondents with poorer health outcomes are significantly more likely to get vaccinated than those with good or excellent health. This is what the odds ratios in Tables 3 and 4 show.

**Table 4.**
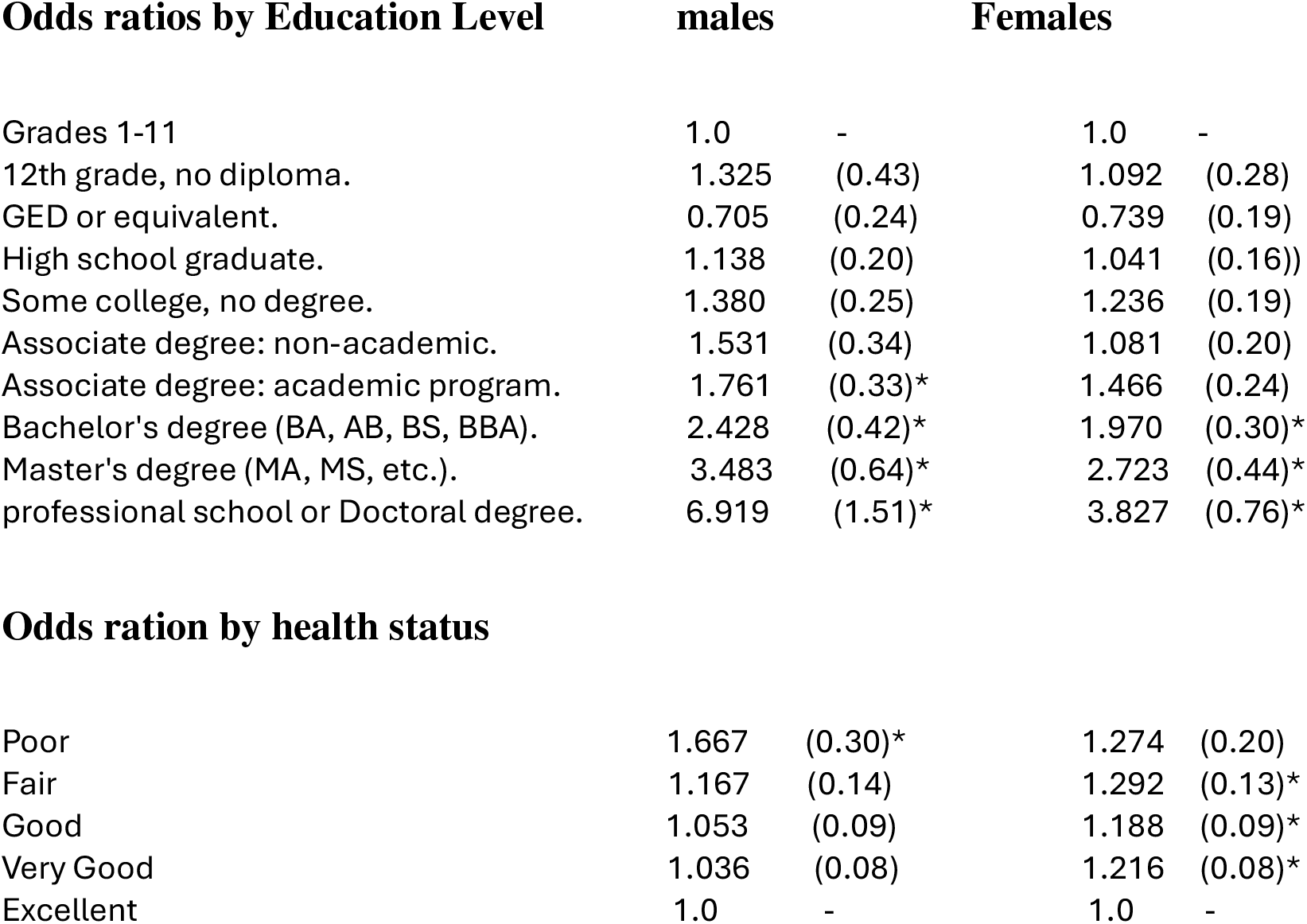
United States Odds ratios by educational level and health status.

**Odds ratios by educational level and health status**
| **Odds ratios by Education Level** | **males** |  | **Females** |  |
| --- | --- | --- | --- | --- |
| Grades 1-11 | 1.0 | - | 1.0 | - |
| 12th grade, no diploma. | 1.325 | (0.43) | 1.092 | (0.28) |
| GED or equivalent. | 0.705 | (0.24) | 0.739 | (0.19) |
| High school graduate. | 1.138 | (0.20) | 1.041 | (0.16)) |
| Some college, no degree. | 1.380 | (0.25) | 1.236 | (0.19) |
| Associate degree: non-academic. | 1.531 | (0.34) | 1.081 | (0.20) |
| Associate degree: academic program. | 1.761 | (0.33)* | 1.466 | (0.24) |
| Bachelor's degree (BA, AB, BS, BBA). | 2.428 | (0.42)* | 1.970 | (0.30)* |
| Master's degree (MA, MS, etc.). | 3.483 | (0.64)* | 2.723 | (0.44)* |
| professional school or Doctoral degree. | 6.919 | (1.51)* | 3.827 | (0.76)* |
| **Odds ration by health status** |  |  |  |  |
| Poor | 1.667 | (0.30)* | 1.274 | (0.20) |
| Fair | 1.167 | (0.14) | 1.292 | (0.13)* |
| Good | 1.053 | (0.09) | 1.188 | (0.09)* |
| Very Good | 1.036 | (0.08) | 1.216 | (0.08)* |
| Excellent | 1.0 | - | 1.0 | - |

The choice of self-reported health is a curious choice for the variable used in measuring the variation in vaccine rates. First, there is a minor problem with the timing of the data collection. Self-reported health is measured at the time of the survey whereas vaccinations have occurred at an earlier time. The Canadian surveys actually give the month in which the respondent received the flu shot and this is usually not the same as the survey date.This, in efect, has the present explaining the past which, from a methodological point of view, is unsatisfactory. It also raises the possibility that the direction of causality may run from vaccination to self-reported health. This implications of this possibility are discussed later in section 4.

There is another problem, but that is related to any variable used to measure the variation in vaccination rates. The vaccination decision for young respondents is often taken by their parents. So what is needed is parent self-reported health, income, or education not the values for the respondent. This is not available in any of the surveys. For education there is another problem and that is for young adults education is not competed education so current education is inappropriate. Fortunately, limiting the sample to ages greater than 25, where most respondents will have completed their schooling, did not alter any of the results.

The most important drawbacks from using self-reported health are, first, it is not the best variable to use in determining variation in vaccination rates. The maximum difference in vaccine rates is quite small for self-reported health about 12 percentage points for Canada an only 4 percentage points for the US. For US education data the difference is much larger at 32 points. The vaccination rate for PhD. holders is more than twice that of respondents with a GED qualification. It is also the case that the responders in the low educational categories have much lower vaccination rates than those in any of the self-reported health categories. As noted earlier, research in this area is supposed to uncover the groups that can be singled out as targets for vaccination programs. Education does this job better than self-reported health.

There is another curious feature associated with the use of self reported health and that is that self reported health is strongly correlated with educational attainment but they have different impacts on vaccination decisions.

Additionally, there is also much more information to be gained about vaccination rates from using education instead of self-reported health. Table 5 shows the relative contributions of each of these two variables in explaining the variation in vaccination rates. The increase in McFadden’s^4^ R^2^ from adding self-reported health to the education dummies is negligeable. Almost all of the variation in vaccination rates is explained by respondent education.

**Table 5.** Model R^2^ Values.

**Model R<sup>2</sup> Values**
| **Model** | **Canada** |  | **United States** |  |
| --- | --- | --- | --- | --- |
|  | **R<sup>2</sup>** |  | **R<sup>2</sup>** |  |
|  | Males | Females | Males | Females |
| Baseline | 0 | 0 | 0 | 0 |
| Education variables | 0.068 | 0.056 | 0.108 | 0.064 |
| Education and reported Health Variables | 0.071 | 0.058 | 0.109 | 0.065 |

In Table 4 the odds ratios show that better educated American respondents are much more likely to have been vaccinated that those with low levels of educational attainment. It is also interesting to note that within the university degree category respondents with more than a bachelor’s degree are much more likely to have been vaccinated. This within category variability for educational attainment was the reason why the NHIS survey was chosen over the BRFSS survey. Using dummy variables produces reliable results only when there is no within category variation. The BRFSS surveys use only one category for university qualifications. This is a specification error since there are significant vaccination rate differences in the sub-categories within the university category. As Breslaw and McIntosh (1999) showed, this is a measurement error problem and it will lead to biases in the computation of odds ratios.

While this relation holds for the United States it does not for Canada. All the Canadian odds ratios in Table 3 for education are not significantly different from unity.

## 4. Discussion

While the results are independent of when the data was collected there are important differences between the two countries. Respondent education matters in the United States but not so in Canada when it comes to vaccination decisions. In Canada the lowest educational category has the highest vaccination rate. Exactly the reverse is true for the United States. A possible reason is that everyone in Canada is covered by a national medicare system. Everyone regardless of income or education can have a family doctor or have access to medical clinics and flu vaccines are free for everyone. In the United States access to non-emergency health care is limited to those with health insurance or sufficient income to pay for hospital or doctor visits. Vaccines are free only for this group. The conjecture here is that access to medical care is related to having information about the benefits of flu vaccinations; many disadvantaged households have little contact with health professionals and are unaware of the importance of vaccines and are possibly put off by their cost. Of course, there are other possible explanations including the fact that the two countries are just different. La et al (2018 p. 434 ) found large differences in vaccination rate across states: South Dakota with 49.5% and Florida with 30.2% but offered no explanation. European countries also differ in their vaccination rates Jemna et al (2022).

The direction of causality was mentioned in section 3. It is difficult to come to any conclusions about causality relation between vaccination decisions and self-reported health but this is not the case for vaccination decisions and education attainments. Education is a predetermined variable and is, therefore, exogenous, and can serve as a legitimate regressor. What may be a problem is that there could be unobserved variables which are correlated with both the vaccination decision and educational attainments and that may lead to omitted variable biases in the parameter estimates. There are procedures available to deal with this problem. The method of instrumental variables, Greene (2007), is one but there are no proper instruments available. As an alternative a procedure developed by Heckman and Singer (1968) was used to solve this problem. There was no evidence to of omitted variable bias in the odds ratio parameter estimates reported in Tables 3 and 4.

> Attention now turns to question of how these results can be used to design more effective vaccination policies. Unfortunately, it is not so clear how convincing vaccine information can be conveyed to those in the lower end of the American educational distribution. The fact that more highly educated individuals choose to be vaccinated suggests that the process of making this decision is quite complex and that anti-vaccination propaganda may be a factor. This suggests that more detailed and frequent TV advertising that explains why it is a good idea to get a flu shot might be effective in promoting higher vaccination rates among the less well educated. To be most effective they should appear in prime time and be associated with sports events like NFL football, NBA basketball, MLB baseball, and NHL hockey which have large numbers of viewers in both countries.

Other policies that are not specifically targeted at poorly educated respondents can still be effective for this group. For example, having annual vaccine clinics each fall in elementary and high schools. In the Canadian provinces of Ontario and Québec school attendance requires vaccines for a number of diseases like diphtheria, tetanus, measles, polio etc. Flu could be added to the list diseases for which a vaccine is required. Failing this, students could be given a note to take home which invites their parents to bring their children to the clinic where all family members can get vaccinated at the same time. In low-income neighborhoods this is likely to attract respondents who ordinarily would not get vaccinated.

Incentives are sometimes effective in changing behavior. Giving a tax credit to individuals who provide proof of an annual vaccination could encourage more people to get vaccinated. This would be more attractive to low-income respondents; however, many low-income households pay no taxes so other payment schemes might be required. How much should the tax credit be? A modest amount would probably work because resistance to flu vaccination largely driven by ignorance and apathy. As the surveys show the reasons why most Canadians do not get vaccinated is because either they did not think it was important or they never got around to doing it.

There is another incentive option. As noted earlier, US health care is provided by the private sector and many Americans have insurance to cover their health care expenses. American health insurers should be encouraged (or forced) to charge a premium to individuals who have not had a flu shot in exactly the same way that smokers have to pay more than non-smokers for their coverage. Insurance companies collect data on health care costs so it will be easy for them to calculate how much more non-vaccinated clients should pay.

Watson and Oancea concluded that ‘more effort is needed from health policy makers to convince individuals with high self-reported health that they too need to receive an annual flu vaccine’. This misses the essential point which is that vaccination rates are too low for all segments of the population. Vaccination policies have to be more broadly based and inclusive to be effective.

## Data Availability

All the data comes from public use files and is freely available to any academic researcher.

## Ethical Statement

No ethical approval was sought since the data used in the analysis was public use data.

## Footnotes

1 Public Health Canada (2023) and CDC (2024).

2 Weir and Gruber (2016 p.354) write ‘Due to the constant antigenic drift of the influenza virus hemagglutinin (HA) and neuraminidase (NA) surface glycoproteins, the vaccines designed to protect against influenza illness must be updated periodically in an effort to match the vaccine strain with the wild-type viruses circulating in a particular seaon’.

3 This data routinely appears in the Canadian Community Health Surveys. Neither the BRFSS nor NHIS has any information on why respondents decide to have a flu shot.

4 McFadden’s R^2^ is the percentage increase in the ln-likelihood function from the baseline model.

## References

Breslaw, J & James McIntosh (1998). Simulated latent variable estimation of models with ordered categorical data. Journal of Econometrics 87, 25–47.

Cantan, Ben, Charles-Edouard Luyt, and Ignacio Martin-Loeches, (2019). Influenza infections and emergent viral infections in intensive care unit. Seminars in respiratory and critical care medicine. Vol. 40. No. 04. Thieme Medical Publishers.

CDC (2024). Mortality and Morbidity Weekly Report. August 29, 2024.

Greene W.H. (2008). Econometric Analysis 6th edition. Pearson-Prentice Hall. Upper Saddle River, New Jersey, USA.

Heckman, James, and Burton Singer (1984). A method for minimizing the impact of distributional assumptions in econometric models for duration data. Econometrica: Journal of the Econometric Society (1984): 271–320.

Jemna, DV., David, M., Bonnal, L. et al., (2024). Socio-economic inequalities in the use of flu vaccination in Europe: a multilevel approach. Health Econ Rev 14, 61 (2024). 10.1186/s13561-024-00535-1

La, Elizabeth M., et al, (2018). An analysis of factors associated with influenza, pneumococcal, Tdap, and herpes zoster vaccine uptake in the US adult population and corresponding inter-state variability. Human vaccines & immunotherapeutics 14.2: 430–441. 10.1080/21645515.2017.1403697.

Mussio, Irene, and Angela CM de Oliveira, (2022). “An (un) healthy social dilemma: a normative messaging field experiment with flu vaccinations.” Health Economics Review 12.1 : 41.

Public Health Agency of Canada (2023). Seasonal Influenza Coverage in Canada, 2022 2023.

Shoar, Saeed, and Daniel M. Musher, (2020). Etiology of community-acquired pneumonia in adults: a systematic review. Pneumonia 12 (2020): 1–10.

Watson, I., & Oancea, S. C. (2020). Does self-rated health status influence receipt of an annual flu vaccination? Preventive medicine, 131. 10.1016/j.ypmed.2019.105949.

Weir, Jerry P., and Marion F. Gruber (2016). An overview of the regulation of influenza vaccines in the United States. Influenza and other respiratory viruses 10.5 (2016): 354–360.

